# Application of Digital Tools and Artificial Intelligence to the Myasthenia Gravis Core Examination

**DOI:** 10.1101/2024.07.19.24310691

**Authors:** Marc Garbey, Quentin Lesport, Helen Girma, Gülṣen Öztosen, Mohammed Abu-Rub, Amanda C. Guidon, Vern Juel, Richard Nowak, Betty Soliven, Inmaculada Aban, Henry J. Kaminski

**Affiliations:** Department of Surgery, George Washington University – School of Medicine & Health Sciences- USA; Care Constitution Corp - Houston, TX, USA; LaSIE UMR-CNRS 7356 University of La Rochelle, France; Department of Neurology & Rehabilitation Medicine, George Washington University – School of Medicine & Health Sciences – USA; Division of Neuromuscular Medicine, Department of Neurology, Massachusetts General Hospital, Harvard Medical School, Boston, 02114 – USA; Department of Neurology, Duke University School of Medicine, Durham, NC USA; Department of Neurology, Yale School of Medicine, New Haven, USA; Department of Neurology, University of Chicago, Chicago, IL USA; Department of Biostatistics, University of Alabama at Birmingham, Birmingham, AL USA

**Keywords:** telehealth, telemedicine, myasthenia gravis, ptosis, diplopia, muscular weakness, deep learning, computer vision, eyes tracking, neurology disease, clinical trial

## Abstract

**Background:** Advances in video image analysis and artificial intelligence provide the opportunity to transform the approach to patient evaluation through objective digital evaluation.

**Objectives:** We assessed ability to quantitate Zoom video recordings of a standardized neurological examination the myasthenia gravis core examination (MG-CE), which had been designed for telemedicine evaluations.

**Methods:** We used Zoom (Zoom Video Communications) videos of patients with myasthenia gravis undergoing the MG-CE. Computer vision in combination with artificial intelligence methods were used to build algorithms to analyze videos with a focus on eye or body motions. For the assessment of examinations involving vocalization, signal processing methods were developed, including natural language processing. A series of algorithms were built that could automatically compute the metrics of the MG-CE.

**Results:** Fifty-one patients with MG with videos recorded twice on separate days and 15 control subjects were assessed once. We were successful in quantitating lid, eye, and arm positions and as well as well as develop respiratory metrics using breath counts. Cheek puff exercise was found to be of limited value for quantitation. Technical limitations included variations in illumination, bandwidth, and recording being done on the examiner side, not the patient.

**Conclusions:** Several aspects of the MG-CE can be quantitated to produce continuous measures via standard Zoom video recordings. Further development of the technology offer the ability for trained, non-physician, health care providers to perform precise examination of patients with MG outside the clinic, including for clinical trials.

**Plain Language Summary:** Advances in video image analysis and artificial intelligence provide the opportunity to transform the approach to patient evaluation. Here, we asked whether video recordings of the typical telemedicine examination for the patient with myasthenia gravis be used to quantitate examination findings? Despite recordings not made for purpose, we were able to develop and apply computer vision and artificial intelligence to Zoom recorded videos to successfully quantitate eye muscle, facial muscle, and limb fatigue. The analysis also pointed out limitations of human assessments of bulbar and respiratory assessments. The neuromuscular examination can be enhanced by advance technologies, which have the promise to improve clinical trial outcome measures as well as standard care.

---

Telemedicine and multi-modal patient monitoring technologies are poised to revolutionize conventional clinical care and enhance the efficiency of clinical trials.^1–3^ These innovations hold particular promise for patients facing barriers to accessing healthcare, such as those with disabilities, limited economic resources, and lack of caregiver support. Furthermore, these obstacles may deter patients from participating in clinical research studies, which require frequent monitoring visits. Individuals with rare diseases may have more pronounced challenges due to the additional hurdles of limited numbers of care centers and trials for rare diseases.^4^ Improving efficiency of clinical trial performance inclusivity are critically important to advance therapeutic development.^5^

Variability in clinical trial outcome measures is well-recognized. Variation can result from ambiguity in metrics themselves as well as differences in how evaluators.^6–8^ Video-based examinations using accessible technology offer opportunities for synchronous or post hoc quantitative analyses. Improved outcome measures and approaches to clinical care are particularly needed for myasthenia gravis (MG) given its rarity, propensity for fluctuation, and benefit for extensive disease-specific monitoring by subspecialists.^7–9^

A standardized examination for MG, specifically tailored for telemedicine use, was developed at the start of the COVID-19 pandemic.^10^ This assessment, known as the MG-Core Examination (MG-CE), was based on the traditional neuromuscular examination performed in clinics and insights gained from outcome measures used in clinical trials for MG. We took advantage of a bank of recorded video sessions to create algorithms aimed at quantifying various aspects of the MG-CE.^11, 12^ Here, we apply and refine our algorithms with a large cohort of MG patients and a diverse control sample. Our approach consistently identifies key examination metrics and we recognize aspects of the MG-CE that are not reliable. Our methods have the potential to enhance the way the MG-CE is conducted and to provide quantitative assessments that were previously unattainable.

## Methods

### Subjects and Video Recording

Subjects had been recruited to undergo standardized examinations by telemedicine to assess the performance of the MG-CE (NCT05917184).^10^ These video recordings were not made for this study’s purpose and truly reflect a standard telemedicine visit. We accessed recordings of subjects that had been performed twice within 7 days except for one patient with 39 days between videos. Each subject had been evaluated by the same neurologist with board certification in neuromuscular medicine. Control subjects were selected with no self-reported physical limitations and a zero score on the MG-Activities of Daily Living.^13^ MG-CE was performed once by a board-certified neurologist. Fifty-one patients with MG and 15 controls participated (Supplementary section).

All participants provided written consent. The patient study was approved by the central institutional review board of MGNet at Duke University and the George Washington University Institutional Review Board. All patients had clinical characteristics of MG and confirmed by serum autoantibody elevations or electrophysiology. The control study was approved by the George Washington University Institutional Review Board.

The present data set allowed us to further train our algorithms^11, 12^ using a broad spectrum of computer vision and signal analysis tools in combination with artificial intelligence (AI) methods. We systematically used an AI transcription tool AssemblyAI (San Francisco, CA) and standard NLP techniques to time stamp patient reports, such as perception of double vision or counting exercises. The Supplement provides details on Methods for each examination and their technical limitations.

## Results

### Quality Review

We found great variability in pixel number of the patient image and lighting conditions across recordings. The number of frames per second was 25 except for five videos running at 30. In order to maintain a standard for quality of data acquisition, a video was removed from analysis when 1) the individual was too close to the camera to obtain a view of the region of interest of the body, 2) illumination or lack of contrast made for poor pixel number, or 3) voice volume was too low to allow speech evaluation. Supplement Table 2 lists the number of videos discarded for analysis for each metric and the Supplement lists the reasons for their elimination.

### Ptosis Evaluation

A sample of 540 images from the video data set of controls were manually evaluated so that anatomical landmarks obtained were within 2 pixels. We were able to compute the distance of the upper lid to the bottom of the iris and the distance from the upper lid to the lower lid. Figure 1 shows results from 11 of 14 controls. Recordings for ptosis assessment had pixel resolutions ranging from 7 to 28. We found one third of the videos to have poor resolution. The 4-fold difference between the lowest and highest resolution of videos was in large part due to variations in the distance between the subject and the camera. The aspect ratio of the eye width versus height varied from 33 to 55 percent which contributed to variability in video acquisition. The noise in the numerical output was at most four percent after filtering.^11^ No control had a greater than 4 percent variation with the exception of those older than 70. Our use of the 70-year-old boundary needs to be evaluated.

**Figure 1.**
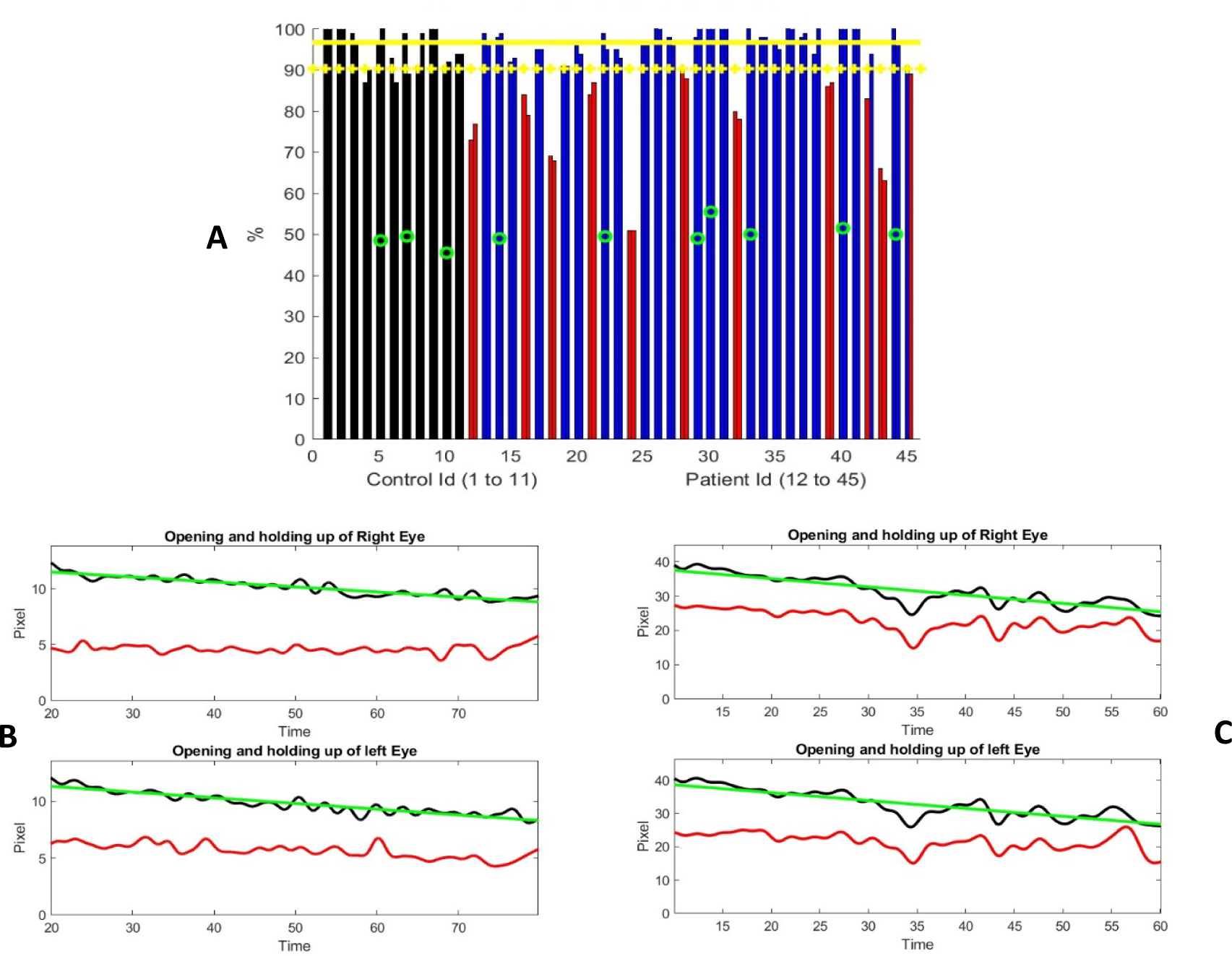
Ptosis Examination. **(A)** Distribution of percentage of change from maximum separation of upper and lower eyelids during the examination. The controls are represented with two adjacent black bars one for each eye. Similarly, patient results are shown in blue or red bars. The most severe ptosis from the two evaluations are presented here. Red columns represent subjects who were significantly different than controls (one standard deviation from the mean). Green circles identify individuals older than 70 years. Yellow horizontal lines represent the mean metric output for control and the mean minus one standard deviation as a threshold to mark the sign of ptosis. **(B, C):** Two examples of the time variation of the distance between the upper lid and lower lid (black curve) and distance from the bottom of the iris to the lower lid (red curve). 1B shows a progressive linear and continuous eye lid fatigue well fitted by a linear square (green curve), while 1C shows measures of a subject struggling to keep the eyes open. Note that the number of pixels marking the eye opening is different for these two subjects, which is representative of the data set, where distance between the subject and the camera is not set the same. Subject in (B) was judged to have moderate ptosis, while subject in (C) was graded from no ptosis to mild and moderate.

Thirty-four patient videos were analyzed (Figure 1A). We found two patterns of lid fatigue. One was a smooth linear drop of the upper lid approximated by the negative slope obtained by linear square fitting. The second was a more chaotic behavior when the patient appears to struggle to maintain upgaze (Figure 1B and 1C). Eleven of 34 patients exhibited lid fatigue. Since we had established a four percent variation for controls, we would not expect false positive results for patients above that threshold. Three patients progressively tilted their head backwards to compensate for ptosis compromising measurement precision, which was detected by the change in vertical dimension from the chin to the top of head. We were able to quantitively and continuously monitor lid fatigue.

### Ocular alignment

Figure 2 shows results from barycentric coordinate determination of the visible iris boundary for 13 controls. The two failures related to subjects turning their heads or poor lighting conditions. We appreciated that eye opening is marginally smaller for controls during this test compared to ptosis assessment. Ocular alignment could be determined within five percent. Therefore, a five percent error is the threshold above which ocular misalignment could be estimated.

**Figure 2.**
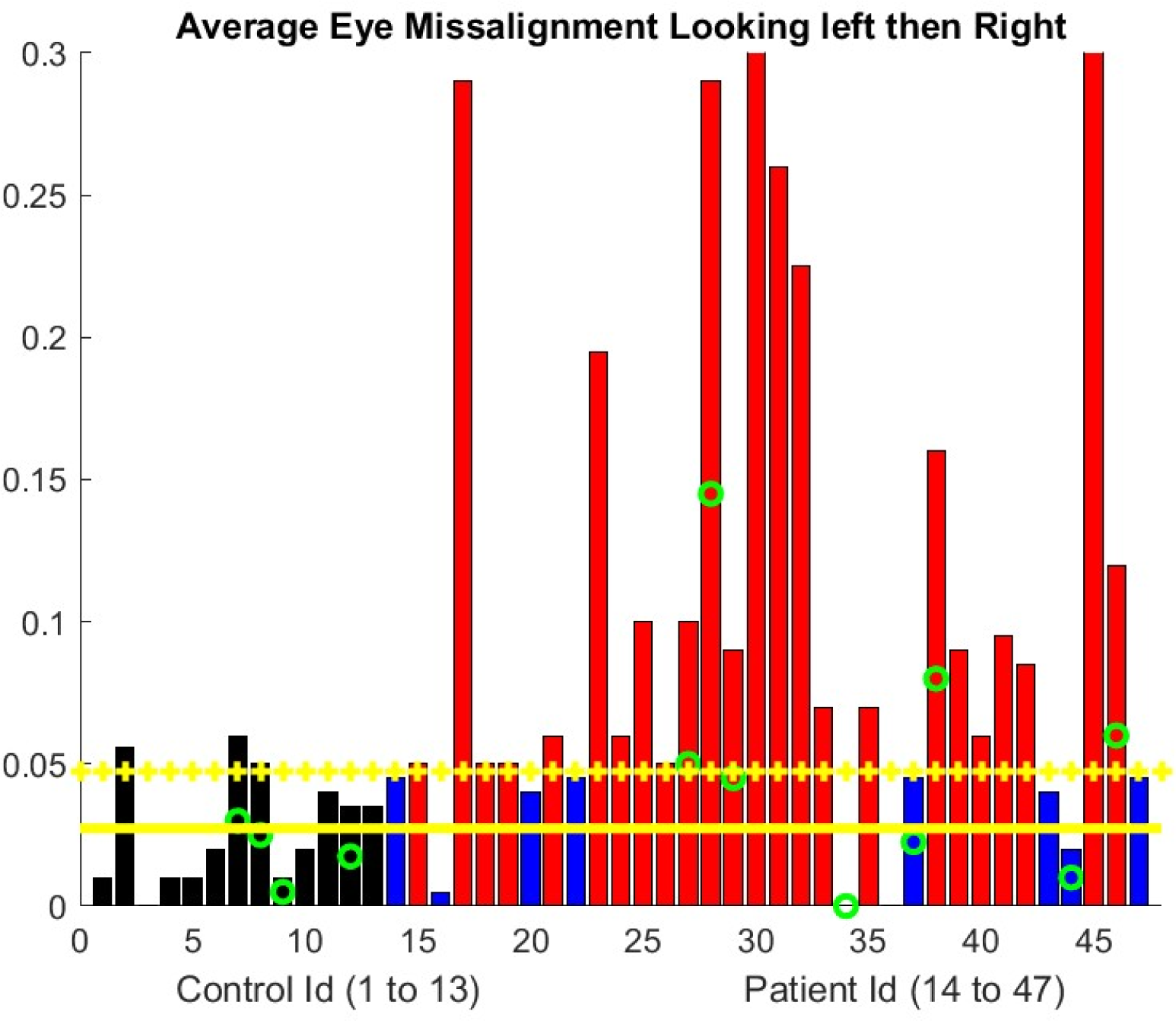
Ocular Alignment. Ocular alignment as assessed by the difference in the barycentric coordinate (0= no misalignment). The progressive misalignment between both eyes uses a least square approximation of the bariatric coordinate change during the exercise. Controls are represented with the black bar. MG patient results are shown with blue bar if there is no significant misalignment developing during the examination and a red bar otherwise. Green circle indicates the subjects are older than 70 years. Yellow horizontal lines represent the mean metric output for control and the mean plus one standard deviation as a threshold to mark the sign of eye misalignment.

For patients the average vertical opening of the lids during eccentric gaze for the diplopia test was less than for controls. We obtained a ratio of 0.63 with a standard deviation of 0.17. Consequently, the number of pixels available to assess the iris position was less than during the ptosis evaluation. The ocular misalignment was computed only along the horizontal.^12^ In two patients, our algorithm failed because the misalignment was primarily in the vertical direction. A few patients had a stable alignment of their eyes with respect to the barycentric coordinate we computed, but still reported double vision, while others had double vision with no misalignment. In fact, the resolution of videos usually does not allow the human examiner to appreciate ocular misalignment. The NLP algorithm allows precise identification of the time stamp of reported double vision. We observed (Figure 2) that roughly half of the patients did have a drift in ocular alignment during the test.

### Arm Extension

Figures 3 and Supplemental Figure 1 illustrate the elapsed time that the participant can maintain shoulder abduction to approximately 90 degrees with the elbows fully extended. Our algorithm measured the angle formed by the arm and torso as well as the vertical variation. Our assessment requires that the patient remained seated for the exercise and for the camera image to show both arms; however, an examiner can evaluate the exercise with only partial view of the arms. The algorithm computes the total time the participant maintains shoulders abducted up to the two-minute duration and the slope of the decay of the angle formed by each arm and the vertical arm position during the elapsed time.

**Figure 3.**
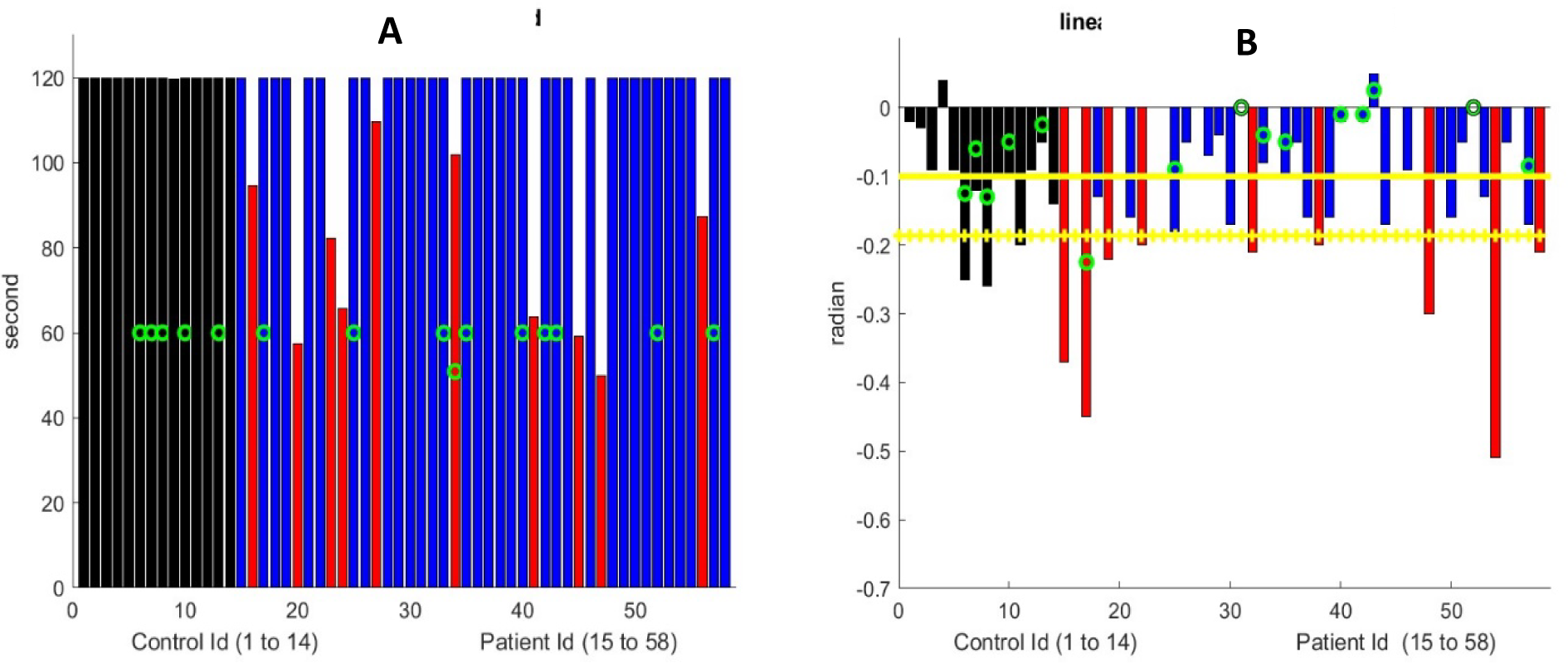
Arm Extension. **(A)** Maximum Arm Extension Time. All controls reached the 120 second’s limit. Nine MG patients marked with red bar could not. The green circle mark patients older than 70 years. **(B)** Linear drift of arm extension in radians for subjects who can extend their arms for 120 seconds. The black bar represents control subjects and blue bar for MG subjects who are similar to controls. Red bar for MG subjects with significantly greater drifts. We show the maximum drift for both arms at both visits for MG patients. Green circle marks subjects older than 70 years. The Yellow horizontal lines represent the mean metric output for control and the mean minus one standard deviation as a threshold to mark the sign of shoulder muscular weakness.

Despite variation in initial arm position among participants, the algorithm successfully analyzed all controls, except one that had an obstructed view of the arms. The average downward drift during the test was −0.05 radian for those younger than 70 years old and −0.15 for those older. Therefore, subtraction of the standard deviation to detect fatigue provides a threshold of −0.12. In other words, any less negative slope would be an indication of fatigue for patients younger than 70 years and −0.24 for those older.

Ten of 44 patients could not hold their arms extended for two minutes, and an additional nine had significant drift (Figure 3, Supplement Figure 2). The decay of that angle is in first approximation linear. In other words, the drift of the arms from the horizontal is a continuous (linear) process, that starts from time zero. We assessed manually that both metrics were computed correctly with great accuracy, provided that the torso, head, and both arms are within the frame of the video. In comparison to the formal MG-CE scoring arm drift is not assessed, only holding for 2 minutes. Quantification of drift by examiners viewing of the video is difficult and may be impossible. However, the digital algorithm, identifies an abnormality.

### Sit-to-Stand

We evaluated the ascending phase elapsed time for subjects who could stand with arms crossed (Figure 4). All controls were able to stand with arms crossed or uncrossed. We discarded five control and nearly half of patient videos because either the head or the hips were not seen. An additional three subjects had only partial view of the head upon standing. The automatic identification of the sit-to-stand time given the time lag of the algorithm and the limited number of frames per second in the video led to poor accuracy (Figure 4). We assessed all entries manually and found an error less than 20 percent for 7 controls and 10 patients for those videos with complete view of the body during the test. This error was 40 percent for the entire data set, i.e. 10 controls and 28 patients for those with a video with partial view of the head in standing position. We easily detected automatically when the patient could not stand.

**Figure 4.**
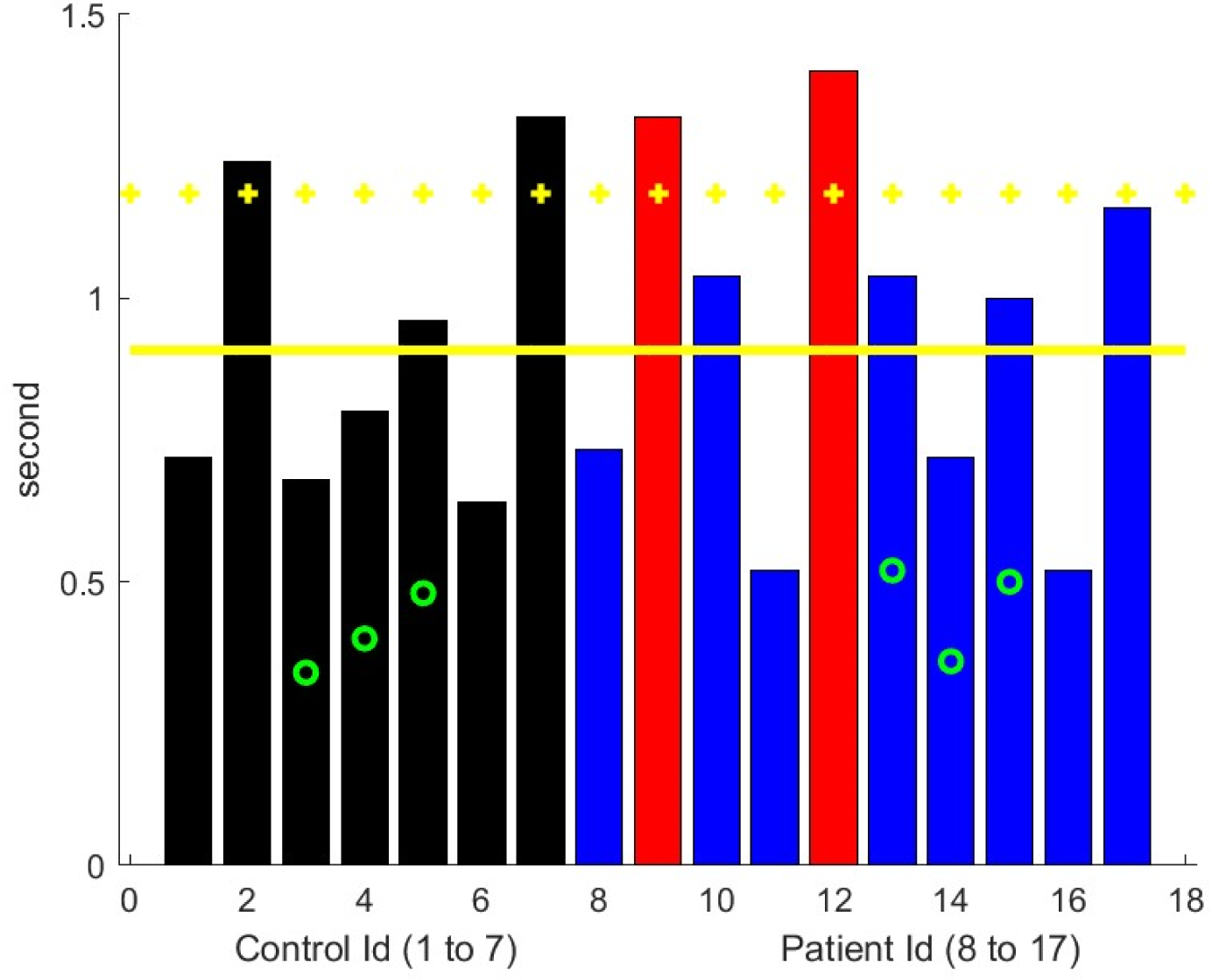
Ascending time of sit-to-stand. Control results are in black and MG subjects in blue with red bars with significantly worse performance than controls. Green circle marks are for patients older than 70 years. The Yellow horizontal lines represent the mean metric output for control and the mean plus one standard deviation as a threshold to mark the sign of leg muscular weakness.

### Count to 50

Using NLP, identification of the sound track time windows that correspond to the exercise is successful with great accuracy. Dysarthria was not evident in controls by the neurologist examiner. We first computed the time length of the count-to-50 for each patient and compared the elapsed time of the two evaluations. We observed a relatively good consistency of the time for the same patient at two different visits, but there was great variability among patients in the frequency of counting with elapsed time varying from 0.5 to 1.5 seconds.

We were able to compute lip and mouth dynamic motion. The acceleration of the mouth motion vertical component could be an indicator of muscle weakness of the lips, and to some extent, cheek and jaw muscles.^14^ Mouth motions of controls greater than 70 years old was generally slower. Four of 49 patients were rated as having dysarthria by physicians, and five with possible dysarthria, based on one of two evaluators identifying the number at which dysarthria appears (publication in preparation). A low acceleration number was present for all these patients (Figure 5). However, weakness of vocal folds, pharyngeal, tongue and soft palate muscles can cause dysarthria. This could be best assessed by sound analysis.^15–17^

**Figure 5.**
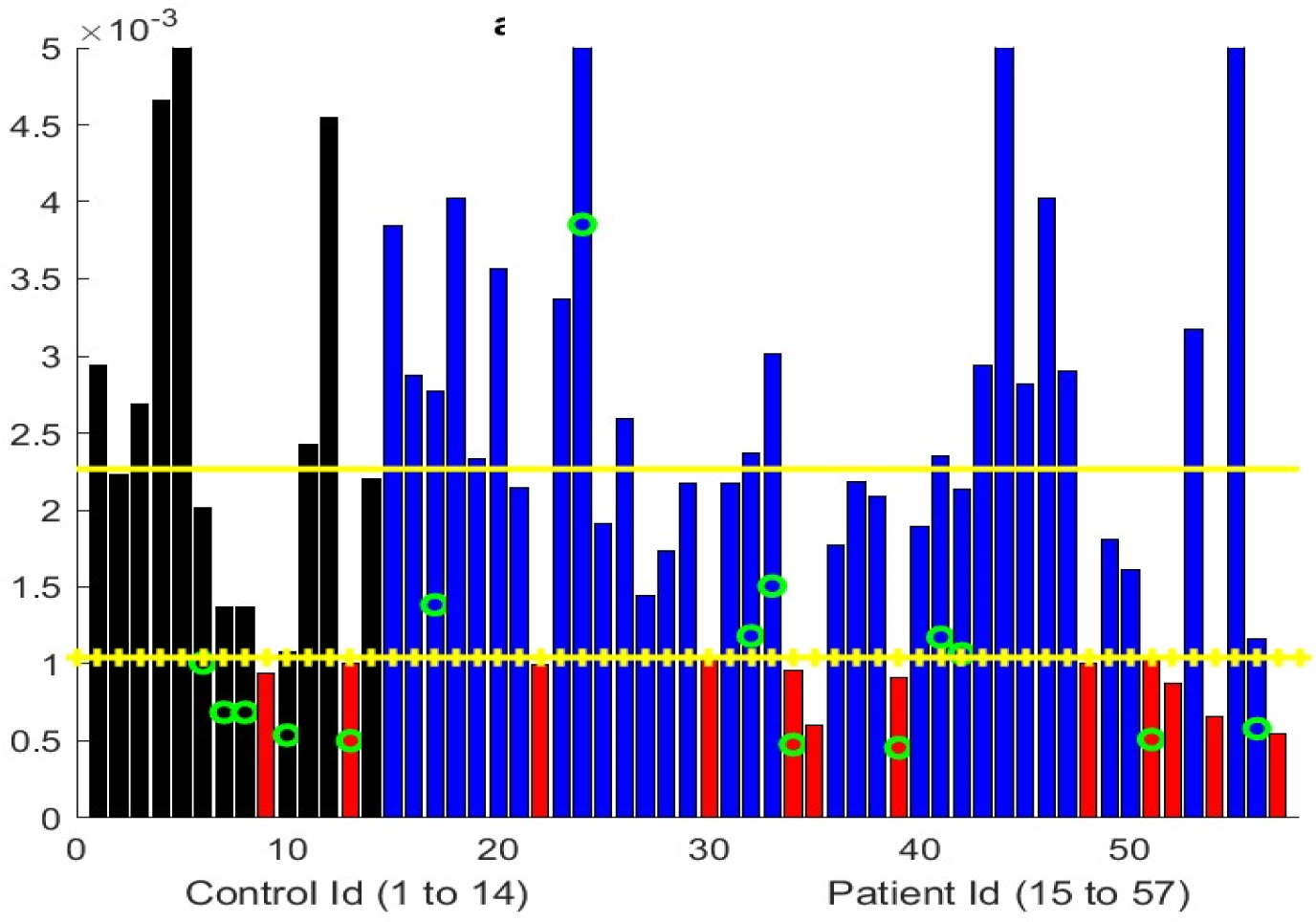
Average variation of the acceleration (normalized distance/second square) of vertical movement of upper lip during counting to 50. Black bars represent controls and MG subjects in blue with red bars identifying significantly worse performance compared to controls. Green circle marks subjects older than 70 years. The Yellow horizontal lines represent the mean metric output for control and the mean minus one standard deviation as a threshold to mark the sign of lip/facial muscular weakness

### Single Breath Count

NLP was able to determine total time for test performance and the number to which the participant counted. All controls younger than 70 years, except one with large body habitus, had no difficulty to count to thirty. We computed the elapsed time for counting on one breath as opposed to the last number said (Supplement, Figure 1). Subjects are not asked to count at a specific rate per the MG-CE instructions and regardless maintaining a particular pace is difficult. We observed that age impacted participants’ performance. However, this measure is likely an underestimate because young controls could have counted for a longer time.

The total time counted varied between visits for individual subjects (Supplemental Figure 2). Instructions state to count as high as possible on one breath, but no attempt at standardization is recommended. It is therefore difficult to use the exercise to judge respiratory function. The variation between visits could be caused by MG, previous activity, or proximity to last dose of pyridostigmine. We were unable to correlate speech features of the count to 50 exercise with the single-breath-count as in our previous study.^11^ This likely related the greater variation across study sites.

### Cheek Puff and Tongue Protrusion

Both evaluations proved to be the most difficult to segment with computer vision. We were able to assess whether the participant was able to form a tight “O” seal of the lips when asked to fill cheeks with air. We computed the pattern/amount of cheek deformation in both the cheek puff and tongue to cheek maneuvers as described previously.^11^ However, variations in anatomy precluded establishing baseline values to compare the control and MG groups.

## Discussion

We have demonstrated that asynchronous quantitative analyses of the MG-CE are feasible. For most exam components, our algorithm distinguished significant differences between the MG and control participants. Contrary to concerns regarding the inferiority of telemedicine examinations compared to in-person assessments,^1, 18^ we found that significant potential exists for quantifying the neuromuscular examination through objective analytical methods.

Despite these videos *not* being originally recorded or optimized for our analyses, a significant majority could still be utilized to derive continuous measures of fatigue in MG patients. The Zoom videos usability was directly proportional to the pixel resolution, uniformity of recording, and optimization of the patient’s lighting and environment. Telehealth offers a unique opportunity to leverage digital technology and AI^3, 19, 20^ as all communication is in digital form and can be archived, revisited, and analyzed with improving algorithms. The manual work associated with this process was nontrivial and involved human review of 50 hours of videos. This is typical of rigorously developed AI applications, which require initial high levels of human input to assure accurate algorithm development.^21^ Digital processing supplemented by AI will make the process more automatic, efficient, and consistent as we optimize the technology.

Our AI algorithms allowed for quantification of the ocular examination, which is not possible during routine clinical assessments by telemedicine or in-person. Our analytics allow an estimate of the marginal reflex (distance form light reflection on pupil to upper lid) from the pre-recorded videos provided that the lighting conditions and pixel resolution were sufficient to see the pupil clearly with respect to upper lid position. Consistent with clinical practice, we did not always detect ocular misalignment, an objective measure, when patients report double vision. Specialized eye movement recording or ophthalmological evaluation, which is not commonly performed by neurologists, is required to definitively assess ocular misalignment. Patients may also exhibit central adaptation to ignore the false image or may have vision impairment in one eye, which limits binocular diplopia. Our algorithm did not assess vertical misalignment, which would have produced double vision. MG patients had a linear drift or chaotic instability of horizontal eye position, most likely indicative of neuromuscular transmission failure.^22^ Most patients demonstrated fatigable ptosis while maintaining a lateral eye position. This made automatic detection of the anatomical landmarks for ocular alignment more challenging to measure in MG patients.

We were able to quantify extended arm position by elapsed time of abduction and downward drift. Drift represents a continuous linear process that can be challenging to track by an examiner, whereas the instability of arm position can persist until patients suddenly drops their arms. These observations may reflect specific aspects of neuromuscular transmission fatigue.^23^ The drift suggests a gradual reduction in the number of active muscle fibers capable of generating sufficient force, while the abrupt drop indicates the simultaneous loss of numerous fibers responsible for maintaining arm position. Both physicians and patients recognize these distinct phenomena. Central or musculoskeletal factors could influence these results.^24, 25^

Standing from a seated position is a complicated movement. Weakness, sensory deficits, pain, and multiple other factors, including compensatory adjustments, may influence the movement. Variations in test performance, most notably the patient’s environment and the examiner’s viewing position, also complicate our assessment. This exercise has the greatest potential for optimization to attain a clinically useful score, especially given the importance of rising from a seated position as a measure of overall function.^26^

We successfully utilized counting-to-50 and single breath counts to develop measures that could not be assessed by an examiner simply viewing the video. No reliable quantification of the deformation of the cheek could be developed for the cheek puff task. Anatomical variation among subjects contributed further to our inability to derive a reliable. The cheek puff and tongue-in-cheek exercise were not suitable for deriving a method to assess. We question the utility of these tasks in the telehealth evaluation since the examiner cannot touch the cheek to evaluate muscle strength.

Despite the limitations of the count-to-50 and single breath count, we were able derive assessments amenable to quantification in a manner different from the exam’s original intent. Lip and jaw movements could be evaluated with great accuracy and consistency across the two videos. Abnormalities were identified for patients judged to have dysarthria by physician examiners. The single breath count is frequently thought to be a good bedside test of respiratory function; however, formal evaluations indicate only mild to moderate correlation with respiratory parameters in patients with less severe weakness.^27, 28^ We found great variation among subjects in counting speed and that elapsed time was a more consistent metric.

The process of digitalization and its utilization of AI introduces new dimensions that are beyond the scope of human perception. Further investigations will define the added value of increased accuracy offered by digital metrics compared to traditional observations as well as the utility of incorporating these new metrics into clinical trials and practice. The digital algorithm outputs require an additional step to transform numerical data into a meaningful score. This process can be likened to converting a laboratory test into a disease progression score, as for example a CD4 count for HIV infection.^29^ Our hypothesis is that digital processing inherently reduces ambiguity and hidden assumptions in protocol tasks, thus potentially enhancing the quality of scoring and provide new metrics to model nervous system function.^30^ Furthermore, we envision our digital framework for conducting the MG-CE examination as an opportunity to (i) enhance physician training before clinical studies, (ii) fully leverage a dataset accumulated during a clinical trial with limited human effort for subsequent analysis, and (iii) facilitate an agile approach to clinical trials, allowing real-time examination of data to identify and address potential gaps and errors in data acquisition as early as possible.

## Data Availability

All data produced in the present study are available upon reasonable request to the authors

## Acknowledgements

Funding: The work was supported in part by the MGNet a member of the Rare Disease Clinical Research Network Consortium (RDCRN) NIH U54 NS115054. Funding support for the DMCC is provided by the National Center for Advancing Translational Sciences (NCATS) and the National Institute of Neurological Disorders and Stroke (NINDS). Additional support was provided by NSF-I Corp award 838792 (47354/1/CCLS 91906F). Dr. Garbey is CEO of Care Constitution, which supported digital tools and AI algorithms.

---

**Supplemental Figure 1.**
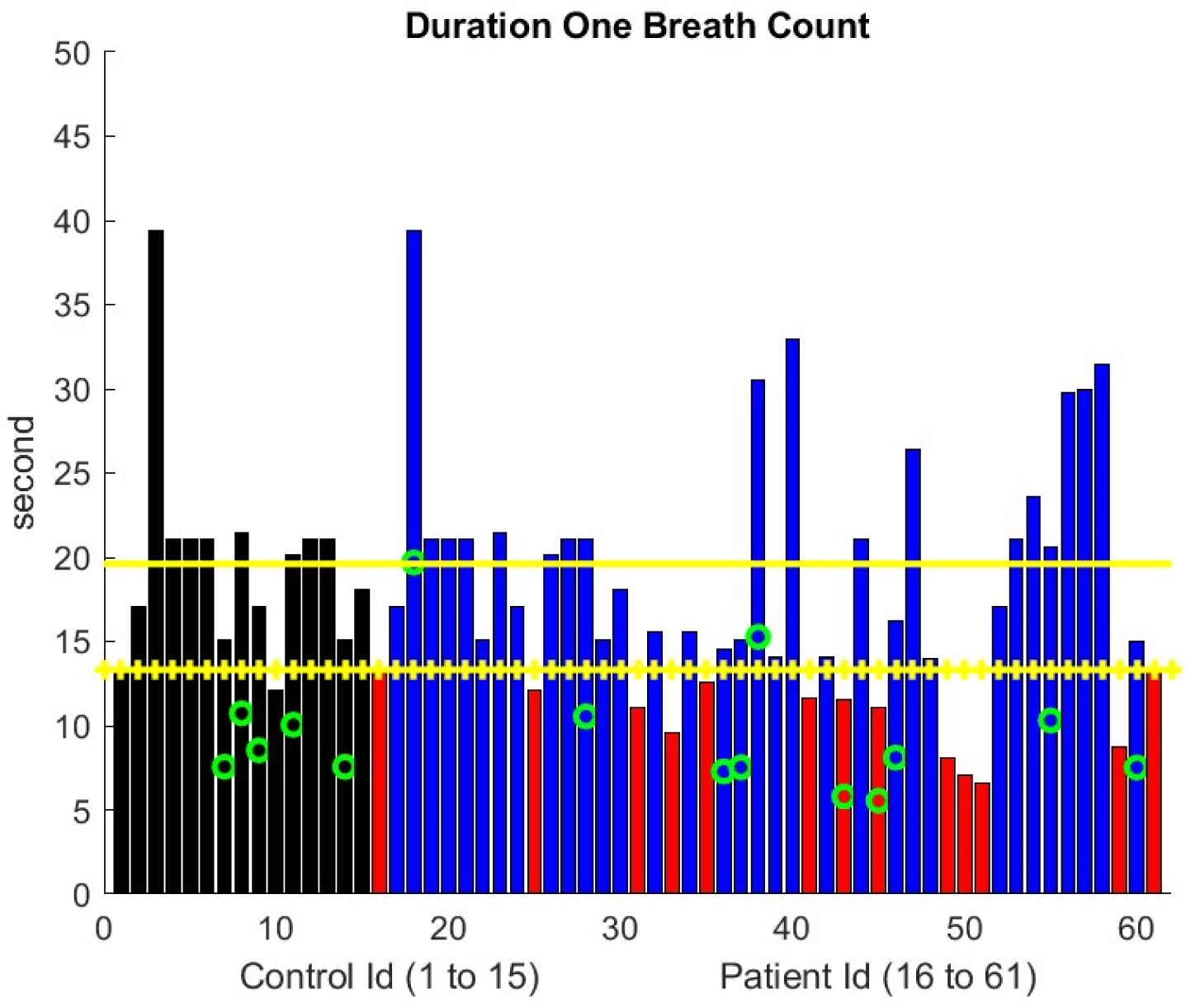
Duration of One Breath Count. Elapsed time for counting on one breath. Control results represented by the black bar and MG subjects in blue except red bars showing significant differences in time of count. Green circle marks subject older than 70 years. The Yellow horizontal lines represent the mean metric output for control and the mean plus one standard deviation as a threshold to mark the sign of breathing weakness.

**Supplement Figure 2.**
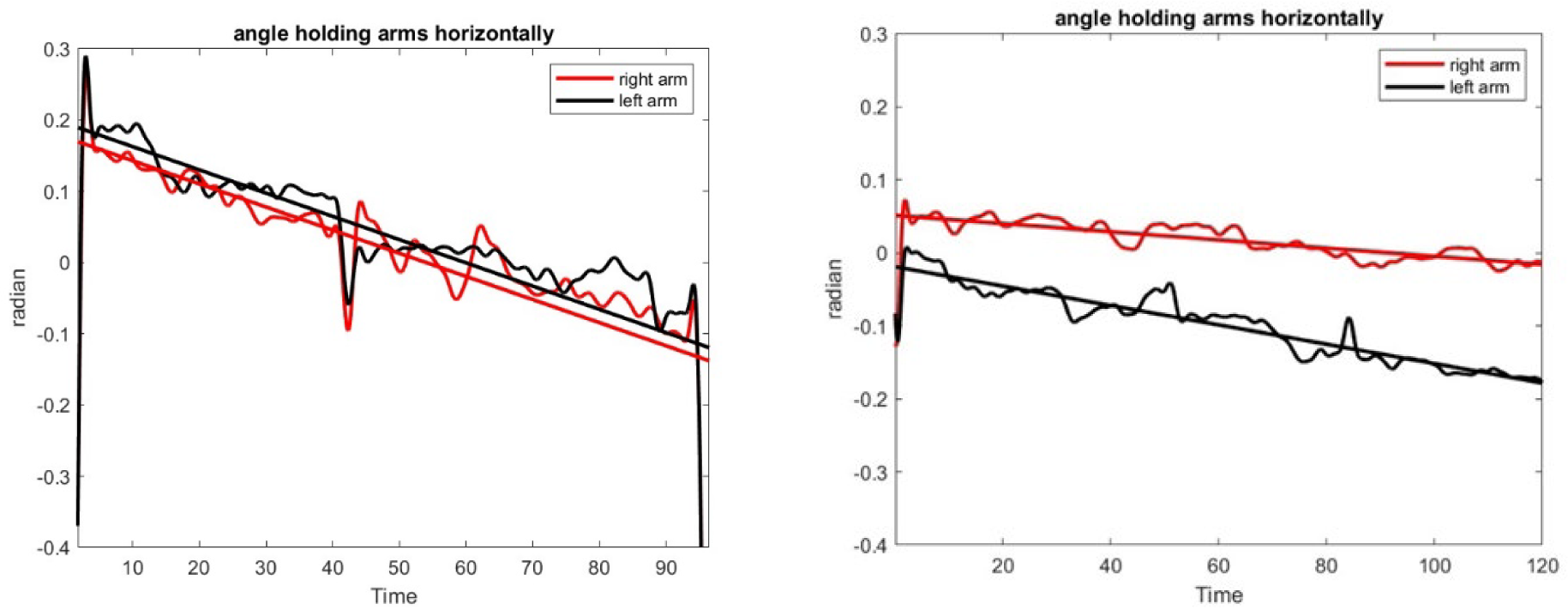
Linear drift of the arms during extension. Example of the linear drift of arms during arm extension. (A) shows a subject with symmetric drift while (B) identifies asymmetric weakness.

